# Health effects of wildfires

**DOI:** 10.1101/2023.04.10.23288198

**Authors:** Carlos F. Gould, Sam Heft-Neal, Mary Prunicki, Juan Antonio Aguilera-Mendoza, Marshall Burke, Kari Nadeau

## Abstract

We review current knowledge on the trends and drivers of global wildfire activity, advances in the measurement of wildfire smoke exposure, and evidence on the health effects of this exposure. We discuss methodological issues in estimating the causal effects of wildfire smoke exposures on health. We conduct a systematic review and meta-analysis of the effects of wildfire smoke exposure on all-cause mortality and respiratory and cardiovascular morbidity. We conclude by highlighting high priority areas for future research, including leveraging recently-developed spatially and temporally resolved wildfire specific ambient air pollution data to improve estimates of the health effects of wildfire smoke exposure.

## 1 INTRODUCTION

In recent years, headlines reading ‘The world is on fire’ have been run in newspapers globally as high-profile wildfires burned in Australia, the Amazon rain forest, Chile, Russia, Portugal, and in the western United States and Canada. Accompanied with striking satellite imagery of wide swaths of the world blanketed in smoke, it seems clear that wildfires negatively impact health: the air is toxic and the blazes destroy property and traumatize communities. And yet, unlike typical ambient air pollution, the impacts of wildfire smoke exposures on health remain incompletely understood. Given that climate change is projected to increase the frequency and size of wildfires in many parts of the world in coming decades, an improved understanding of the health impacts of wildfire is an urgent public health priority.

The purpose of this review is to discuss current knowledge on the trends and drivers of global wildfire activity, techniques and recent advances in the the measurement of wildfire smoke exposure, and available evidence on the myriad health effects of wildfire smoke exposure. We define wildfires as uncontrolled fires that occur in a natural environment, such as forests, grasslands, or prairies. These fires can have both proximate and distant direct and indirect health impacts, ranging from injury from fires, heat, and property damage to respiratory impacts from smoke inhalation to trauma-related mental health harm. We critically review empirical methods for assessing the health effects of wildfire smoke exposure, offering suggestions for future studies to improve methodological consistency and rigor. We then quantitatively synthesize existing literature on the effects of ambient wildfire smoke in a meta-analysis on same-day all-cause mortality, respiratory related emergency department (ED) visits and hospitalizations, and cardiovascular related ED visits and hospitalizations. We conclude by commenting on existing gaps in our knowledge of the health effects of wildfires and potential solutions for addressing their impacts.

## 2 TRENDS IN WILDFIRE ACTIVITY AND DRIVERS

Wildfire activity is driven by a complex combination of climate, ecological, and human factors. Available evidence suggests climate-related factors like temperature and precipitation are the most important drivers of large-scale patterns of wildfire activity.^1^ Climate induced warming and drying has led to increases in the frequency and severity of fire conducive weather conditions is nearly all global regions over at least the last fifty years.^1–4^ A growing population, particularly in the wildland urban interface (WUI), can also lead to more frequent human-caused ignitions; in the future, climate-induced changes in weather systems may also increase the frequency of natural wildfire ignition sources such as lightning.^5^ This combination of factors is likely to generate widespread risk of increasing wildfire activity in the coming decades.^1, 3, 6, 7^

Despite these trends in climate patterns, observed trends in wildfire burned area vary across regions and global average burned area actually declined between 2000 and 2020,^4^ in large part driven by human factors, such as land use transition, forest management, and other land management practices.^4, 8^ In North America, forest management practices and decades of fire suppression have led to an abundance of fuel (i.e., vegetation) which, due to increased summer temperatures and extreme drought, has become more arid and flammable over time, leading to a sharp increase in fire activity.^6, 9^ Other fire-prone regions have seen less clear trends in fire activity. In Australia, levels of fire activity have been steady since 2000,^2, 4^ while changing land use practices in parts of South America and Africa have led to declines in fire activity since 2000.^4^ Irrespective of longer-term trends, fire activity is highly variable year-to-year, and extreme fire activity years are observed even in regions with decreasing trends.

## 3 DIRECT HEALTH EFFECTS OF WILDFIRES

### 3.1 Firefighters

Those that fight fires to limit fire spread often put themselves directly in harm’s way. There are a range of well-documented occupationally-related short-term health risks and an emerging literature on long-term health impacts among front-line wildland firefighters.^10–12^ Wildland firefighters face multiple health hazards, including exposure to smoke, intense heat, low oxygen conditions, excess noise, physical hazards like falling trees, burning debris, and ash, and long working hours with minimal rest and protections.^10, 11^ These hazards have been directly associated with a range of negative health outcomes, including poorer respiratory health, cardiovascular health, mental health, dehydration and malnutrition, and acute physical injuries.^10–12^ To establish these short- and medium-term associations, studies often document within-subject changes in health outcomes before and after work shifts or across fire seasons. For example, Gaughan et al.^13^ found declined one-second forced expiratory volume — a measure of acute lung function — after work shifts among 17 firefighters. Long-term health impacts from occupational exposures among wildland firefighters are less well-documented, though there is an emerging literature suggesting elevated risks for cancer,^14^ cardiovascular disease,^15^ and biomarkers of aging.^16^

### 3.2 Nearby communities

Wildfires can affect health and well-being in local communities through a variety of channels, including physical damage to homes and infrastructure, air quality degradation, loss of livelihoods and income, and disruption of local ecosystems.**^?^**^, 17, 18^ In particular, studies have documented a range of negative mental health outcomes during and after wildfire events in local and distant communities, including elevated rates of post-traumatic stress disorder,^19^ depression,^20^ anxiety,^21^ and substance use.^19^ Wildfires can impact mental health through a variety of pathways at the individual, social and community level, in living and work conditions, and at the ecological level; for example, through reduced sleep duration and quality, reduced physical activity, increased perceptions of risk and anxiety, isolation from others, forced evacuations and/or relocations, reduced access to livelihoods, and loss of nature.^18, 22–24^ Recent work has emphasized the need for broad measures of social impacts from wildfires — as opposed to more common outcomes like buildings burned and resources spent on suppression efforts — that provide more comprehensive pictures of the impact of wildfires on community health and well-being.^17^

Given the vulnerability of some communities to impacts of environmental hazards,^25, 26^ community resilience can be considered a critical factor in mitigating the health effects of wildfires.^27–29^ Beyond the basic conceptualization of resilience, i.e., the ability of a community to prepare for, respond to, and recover from natural disasters or other types of hazards, recent work has advanced the concept of adaptive and transformed social-ecological resilience.^29^ In this framework, communities and social systems adapt to new dynamics (i.e., increased wildfire activity) by changing aspects of the system (e.g., land-use planning) and intentionally transform to acknowledge the changing role of fire in social-ecological systems such that the design of systems is future facing.

We increasingly live in wildfire-prone regions, so it is becoming more and more important to develop and implement policies that anticipate and plan for fires — as we do for other natural hazards like floods, earth-quakes, and hurricanes.^27^ Such strategies include better mapping localized fire hazards, adopting land-use and zoning regulations to limit development in the most fire-prone areas, implementing appropriate vegetation management practices, and evaluating planning and wildfire warning systems.^27, 29, 30^

## 4 WILDFIRE SMOKE

### 4.1 Composition

Wildfires emit a mixture of particles and gaseous pollutants that are known to negatively impact human health, including particulate matter (PM), carbon monoxide, nitrogen oxides, and volatile organic compounds.^22, 31–33^ Ground-level ozone can form secondarily through photochemical reactions. Depending on the materials burned, heavy metals like lead and mercury can also be emitted. Furthermore, wildfire smoke has been documented to contain toxic carcinogens – not unlike cigarette smoke – such as benzene, benzo[a]pyrene, and dibenz[a,h]anthracene.

Wildfire-specific PM likely has a different toxicological profile from PM originating from other sources;^31, 33^ however, the relative toxicity of wildfire derived PM compared to PM from other sources remains uncertain. The amount and composition of pollution emitted from a specific fire varies depending on the fire’s size, temperature of combustion, materials burned (e.g., grasses, tree species, buildings, vehicles), distance the smoke has traveled, and environmental conditions like wind speed, temperature, and humidity.^32–34^

### 4.2 Measurement

To enable studies focused on the health impacts of wildfires, approaches to estimating wildfire smoke exposures must be able to both separate wildfire smoke from other pollution sources and estimate exposures everywhere people live in a temporally and spatially disaggregated manner. Multiple approaches to estimating wildfire-specific ambient air pollution concentrations and exposures have been employed in the literature, each with strengths and limitations.^35–37^

Ground monitors can provide the most accurate estimates of surface pollutant concentrations, but they do not distinguish pollution from wildfires from that from other sources and they are sparsely located in most regions.^38, 39^ Aside from ground monitors, studies have employed atmospheric chemical transport models (CTMs),^40, 41^ dispersion models, and statistical models including machine learning approaches^40, 42–44^ to estimate ambient wildfire smoke concentrations.

CTMs are complex numerical models that simulate atmospheric chemistry dynamics. By directly modeling the movement and evolution of wildfire emissions, they can be used to estimate local pollution concentrations attributable to wildfires. Frequently, CTMs are run with and without emissions from fires to estimate wildfire-specific pollution concentrations. CTMs can characterize complex chemical processes that drive pollution transport, but are computationally intensive and thus hard to run for large spatial areas at high resolution over meaningful time scales; they are also sensitive to uncertain inputs (e.g., emissions from a given fire).^45, 46^ Dispersion models use meteorology and simplified physics to model the transport of pollution emissions and are less computationally intensive than CTMs but may fail to capture some of the complex processes modeled in CTMs.

In contrast to CTMs and dispersion models, statistical models do not attempt to model atmospheric chemistry and instead characterize the direct relationship between wildfires (frequently, the presence of wildfire plumes) and surface pollution concentrations measured at ground monitors. They typically incorporate data from remote sensing measurements of atmospheric aerosols, meteorological conditions, and other measured and modeled data sources that influence wildfire smoke pollution. Because ground monitor pollutant measurements cannot easily be disaggregated by emission source, these models are typically trained on imperfect proxies for surface-level wildfire smoke pollution. Hybrid approaches incorporating outputs from multiple modeling frameworks (i.e., CTMs used as an input into statistical models) have also become increasingly common.^40, 47, 48^

Recently, high-resolution wildfire-specific surface PM_2.5_ concentration estimates have been developed and are publicly available for California^43^ and the contiguous United States.^40, 42^ Daily estimates of total PM_2.5_ that incorporate, but do not distinguish wildfire-specific PM_2.5_, are available at various spatial and temporal resolutions, including for the Western^48^ and contiguous United States^49^ at the daily level, in parts of Canada at the hourly^50^ and daily level,^51^ and globally^52^ at the monthly level. While the choice of wildfire smoke metric may be dictated by availability, existing evidence suggests that this choice can influence the estimated associations between wildfire smoke and health outcomes.^53, 54^ See Table S1 for a summary of available data products and approaches.

### 4.3 Trends

Due to the methodological challenges discussed above, reliable data on global trends in wildfire-specific air pollution are not available. Furthermore, the distinction between wildfires and other types of landscape fire (e.g., crop burning, fire associated with land cover change) is unclear in many regions. Available estimates of global air quality impacts associated with all landscape fire suggest that fire is responsible for about 14% of ambient PM_2.5_ in Africa, 9% in South America, 7% in North America, 4% in Asia, and 2% in Europe in recent years.^55^ However, regional averages mask substantial temporal and geographic variation. For instance, fires are estimated to contribute 61% of total PM_2.5_ in Laos and 45% in Democratic Republic of the Congo, but <1% of PM_2.5_ in Afghanistan. Collectively, 43 million people live in areas where the air quality is ‘unhealthy’ (PM_2.5_ >55 *μgm*^−3^) at least one day per year as a direct result of pollution from landscape fires.^55^

In North America, where air quality monitoring is more comprehensive, evidence suggests that wildfires have played an increasingly important role in determining overall air quality levels over the last several decades.^56^ While wildfires’ impact on US air quality has historically been limited to the Pacific North-west,^40, 57^ smoke’s impact on surface average and extreme PM_2.5_ concentrations is now observed throughout much of the Western US^42, 58^ and as far away as the East Coast.^59^ In 2020 alone, an estimated more than 25 million people in the US were exposed to at least one day with wildfire PM_2.5_ > 100 *μgm*^−3^,^42^ more than three times health-based daily air quality guidelines.^60^

## 5 SUMMARY OF HEALTH IMPACTS OF WILDFIRE SMOKE EXPOSURE

Similar to air pollution from other sources, observational evidence has linked exposure to wildfire smoke with a wide range of human health outcomes (Figure 1) and has been the subject of previous reviews.^22, 32, 61–65^

**Figure 1:**
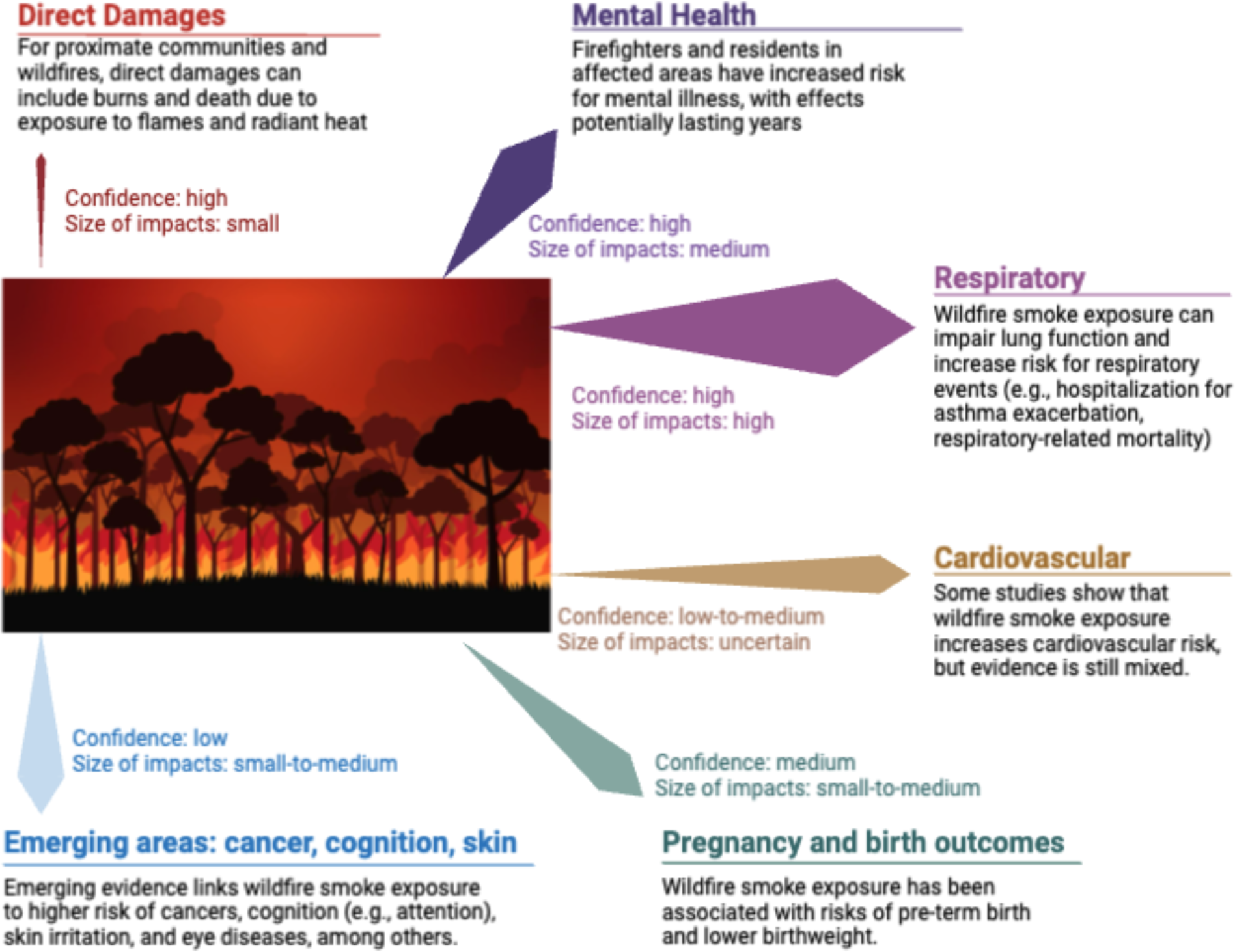
Summary of the health impacts of wildfires.

The literature on wildfire smoke’s impacts on respiratory conditions is particularly rich, showing increased respiratory-related mortality, increased respiratory-related hospitalizations and ED visits, declines in lung function, the exacerbation of chronic respiratory conditions and asthma,^66^ more ambulance dispatches for respiratory-related conditions,^67^ and increased respiratory medication dispensations (e.g., salbutomol).^12^ Additionally, studies have established a link between wildfire smoke and COVID-19,^68^ supported by similar evidence of a relationship between total air pollution and COVID-19 infection susceptibility and severity, as well as other respiratory tract infections.^69^

Despite strong established links between all-source PM and cardiovascular and cerebrovascular health, evidence related to wildfire smoke exposure has been mixed.^70^ Some studies have reported positive associations between wildfire pollution and cardiovascular mortality^71^ and morbidity,^67, 72, 73^ including out of hospital heart attacks,^74, 75^ especially among older adults.^76^ However, a number of studies have also reported non-meaningful association, non-statistically-significant differences, or even declines in cardiovascular-related healthcare utilization (e.g.,^54, 77–80^).

A growing number of studies have identified an association between wildfire smoke and adverse pregnancy and birth outcomes, namely pre-term birth, low birthweight, and birth defects.^81–87^ The causal mechanisms for these relationships are likely to be complex and may encompass both the effect of exposure to wildfire smoke and maternal stress associated with wildfire occurrence.

Recent literature has documented worsened cognitive outcomes,^88, 89^ declines in mental health,^24^ and increased ED visits for headaches^90^ associated with exposure to wildfire smoke. For example, Cleland et al.^88^ assessed cognitive performance among adults and found that wildfire smoke plumes was negatively associated with the estimated attention score on the same day and one week later. Additionally, limited, but growing research that parallels the broader air pollution literature, posits an association between wildfire smoke and neurodegenerative illnesses, like Alzheimer’s.^91^

Emerging literature also links wildfire smoke exposure with skin diseases,^92^ eye conditions^93^ and cancer.^94, 95^ For example, Fadadu et al.^92^ document increased rates of clinic visits for atopic dermatitis or itch in San Francisco during the California Camp Fire in November 2018.

Environmental epidemiological and toxicological studies have investigated the biological mechanisms through which wildfire smoke exposure negatively impact health (e.g.,^96–98^). It is likely that the mechanisms parallel those established in the broader air pollution literature, namely: oxidative stress and inflammation, impaired nervous system function, vascular dysfunction, direct damage when particulates and chemicals enter the bloodstream, and epigenetic alterations, among others.^33, 70, 99, 100^

## 6 CRITICAL REVIEW OF METHODS FOR ASSESSING HEALTH EFFECTS OF WILDFIRE SMOKE EXPOSURE

While there is high biological plausibility and a wide array of evidence that supports the negative health effects of wildfire smoke exposure, estimating the effects of exposure to a given unit of wildfire smoke is challenging. The central empirical challenge, as in other environmental health settings, is in isolating variation in wildfire smoke exposure from variation in other correlated factors that could also affect health outcomes. Absent this ability, measured smoke-health linkages are associational and cannot reliably inform quantitative estimates of the overall health burden of smoke exposure. However, compared to most other sources of variation in air pollution, plausibly random temporal variation in wildfire smoke (i.e., unlikely to be correlated with confounders) offers unique opportunities to separate pollution exposure from other sources of correlated health risk and thus establish plausibly causal concentration-response relationships. However, given difficulty in measuring smoke exposure at broad temporal and spatial scale, these opportunities have not always been exploited in the existing literature.

For instance, one common approach to quantifying smoke-health relationships has been to relate spatial or spatiotemporal variation in health outcomes to similar variation in wildfire smoke at a chosen spatial scale (e.g., zip code), and to adjust directly for variables (e.g., income) that could be correlated with both differences in average smoke exposure and in average health outcomes across locations. The challenge with this approach is that average smoke exposure is correlated in a statistically significant way with a very large set of measurable covariates; see Figure S1 for the census-tract-level correlation between average smoke PM_2.5_ and a small subset of socioeconomic and demographic variables taken from the American Community Survey. Controlling completely for these measured covariates, and for the plausible set of additional unmeasured covariates also correlated with both smoke and health outcomes, becomes exceedingly challenging. As a result, these “regression adjustment” approaches are unlikely to reliably isolate the causal effect of wildfire smoke on health. With that said, advances in biostatistical and environmental epidemiological causal inference methods using high-dimensional matching on observables (e.g., g-computation, generalized propensity score matching) can use these covariates to improve upon standard regression adjustment.

Rather than attempting regression adjustment, alternative approaches have instead utilized temporal variation in smoke exposure, comparing individuals to themselves (as in a case-crossover design) or locations to themselves (as in a time series or panel fixed effects design) over time as smoke concentrations fluctuate. Conditional on seasonal controls and longer-term time trends, such temporal variation is plausibly random — an idiosyncratic function of where exactly a given fire starts and which way the smoke is blown. These designs can best measure the causal effect of short-term variation in smoke (sub-daily to annual), periods over which temporal variation is plausibly random. As desired exposure windows get longer (multiple years or longer), temporal variation is reduced and these designs become more challenging.

A second important issue is in attending to and accurately estimating the potential non-linear shape of the smoke-health concentration-response curve. Recent work shows striking non-linearities in the responsiveness of emergency department visits to daily wildfire smoke exposure,^101^ with increases in total visits at moderate exposures and substantial decreases at high exposures — the latter likely a result of behavioral changes during extreme exposures, such as reduced driving and traffic accidents, that reduce non-respiratory morbidity. Failure to account for these potential nonlinearities, e.g., by estimating a linear concentration-response function, could lead to inaccurate assessments of the overall contribution of smoke exposure to a given health outcome, particularly as increasingly extreme wildfires generate increasingly extreme levels of smoke exposure. Fortunately, given the increasing availability of granular measures of wildfire smoke exposure, estimation of non-linear concentration-response functions is relatively straight-forward. Two approaches are to model health outcomes as a higher order polynomial or spline function of smoke, or to estimate non-parametric “binned” models that model health outcomes as a function of accumulated exposure in prescribed exposure bins (e.g., 0-5 *μgm*^−3^, 5-10 *μgm*^−3^, etc).

A third critical issue is in adequately accounting for the possibility of temporal lags between exposure and outcome. These lagged effects could amplify the total effect of a given smoke exposure, for instance if smoke exacerbates a respiratory infection that leads to an emergency department visit days after the exposure, or they could lead to offsetting effects, for instance by accelerating the speed with which a respiratory infection requires a hospital visit but not increasing the total number of hospital visits; this latter phenomenon, in which contemporaneous and lagged exposures have opposite signs, is often referred to as “displacement” or “harvesting” in the literature. Ex ante, it is unknowable whether either amplification or displacement (or some combination of the two) is occurring. The standard approach to calculating the cumulative (time-integrated) effect of a given exposure increase is to estimate “distributed lag” models, where the health outcome is modeled as a function of contemporaneous and temporally lagged values of wildfire smoke exposure. The cumulative effect of a given wildfire smoke exposure is then correctly calculated as the sum of effects across the contemporaneous and lagged variables; estimating a distributed lag model but not summing the coefficients, as is sometimes done in the literature, does not yield consistent estimates of the total effect of an increase in smoke exposure.

To illustrate the importance of these two concerns — nonlinear effects and lags — we revisit earlier work^101^ and estimate the effect of daily wildfire smoke exposure on emergency department visits, using the universe of cause-coded ED visits aggregated to the zipcode level in California from 2006–2017 and gridded daily estimates of ambient wildfire PM_2.5_.∗ We fit panel fixed effect regression models that include zipcode, day-of-week, county by month-of-year, and wildfire-season by year dummies to control for location and time trending unobservables. In a linear model, the relationship between smoke exposure and ED visits is negative and would lead us to estimate that a total of 1,300 ED visits per year were *averted* attributable to wildfire PM_2.5_ across California 2006–2017 (Fig 2a). In contrast, a nonlinear model (a 4^th^ degree polynomial) indicates that, at low to medium levels (< 25 *μgm*^−3^), ED visits increase, but ED visits decline at higher concentrations. These declines in total ED visits at high daily smoke levels are driven by protective behavior that reduces, among other things, accidental injuries.^101^ Because most wildfire smoke days have low to medium PM_2.5_ concentrations, using the quartic model we estimate an *excess* of 3,300 ED visits per year. When examining respiratory diseases, we can see that the combined respiratory ED visits model is a combination of asthma-related ED visits and respiratory tract infections, which respond differently to increasing wildfire PM_2.5_ (Fig 2b).

**Figure 2:**
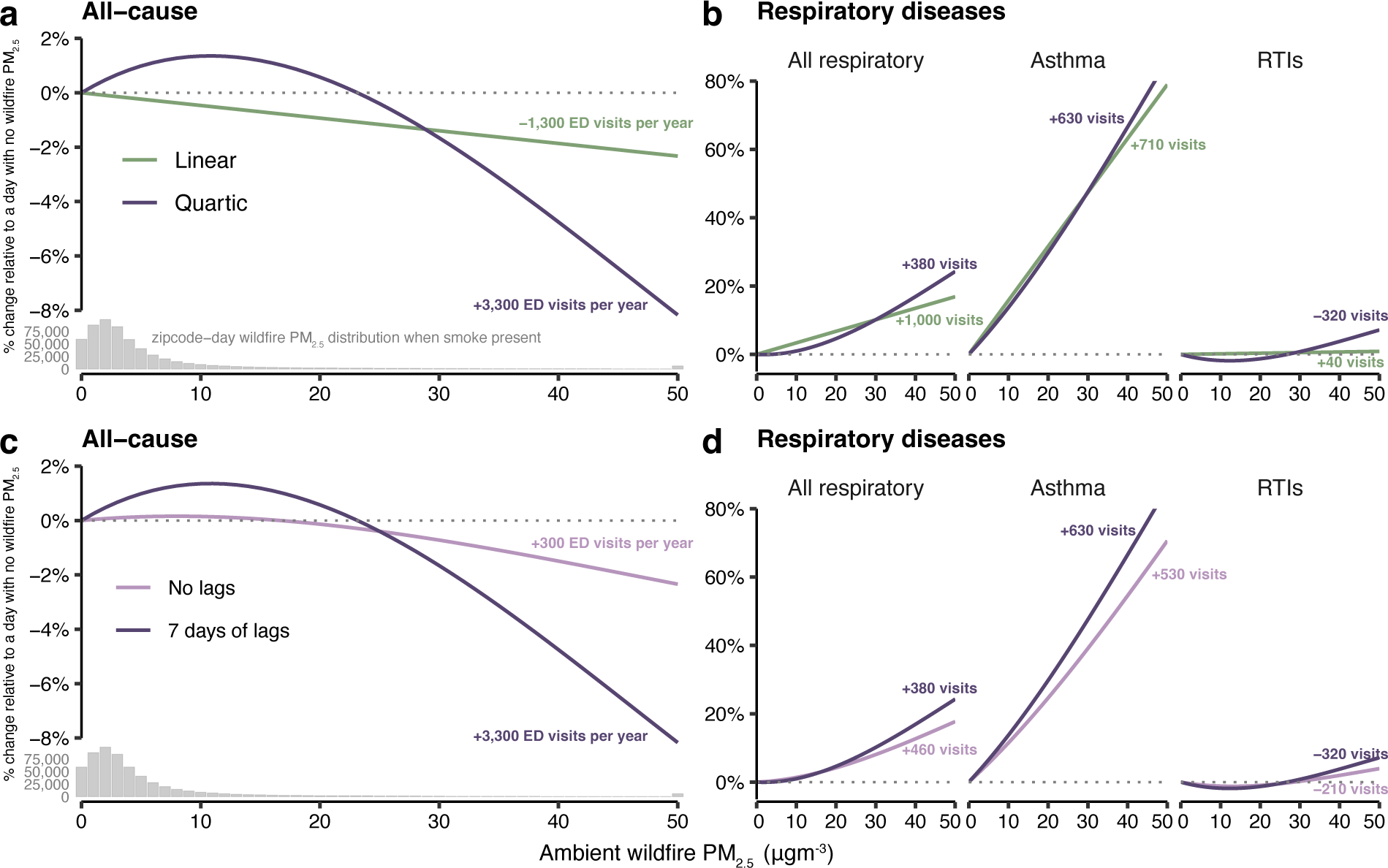
Differing response of daily zipcode level emergency department visit rates to wildfire PM_2.5_ **concentrations according to modeling linear and nonlinear effects and lagged effects in California 2006–2017.** Annotations indicate the total number of ED visits to public facilities attributable to wildfire smoke in California from 2006–2017, with positive indicating excess ED visits and negatives indicating averted ED visits. Histograms in panels a and c indicate the distribution of zipcode-smoke days on days with any wildfire smoke in that zipcode. RTIs = respiratory tract infections.

Failure to account for lags also alters inference of the total effect of wildfire PM_2.5_ on changes to ED visit rates (Fig 2c-d). Accounting for lagged impacts increases the estimated effect of low-to-medium concentrations and also enhances the declines at higher concentrations, with the net effect of increasing excess ED visits from 300 to 3,300 per year.

We use data from Heft-Neal et al. to illustrate these nonlinear and lagged effects, but they have been observed in other studies. For example, several studies have found lower healthcare utilization across a range of outcomes during wildfire periods or in association with increased wildfire smoke (e.g.,^77, 80, 102, 103^).

∗This work was approved by the Stanford University Institutional Review Board and the California State Committee for the Protection of Human Subjects (IRB 2018-255).

However, since most studies seek to examine physiological health responses to wildfire smoke exposures, observed declines in healthcare utilization — an initially counterintuitive finding — are often not explicitly discussed perhaps because they may reflect behavioral rather than physiological responses. The net welfare impacts of these nonlinearities are as yet uncertain and are an area rich for discussion.

## 7 META-ANALYSIS OF EFFECTS OF WILDFIRE SMOKE EXPOSURE ON MORTALITY AND RESPIRATORY AND CARDIOVASCULAR MORBIDITY

With the objective of reliably estimating the extent to which ambient wildfire smoke affects health outcomes, we focus our attention on studies with research designs and statistical methods that use variation in smoke exposure that is plausibly uncorrelated with other drivers of health risk.

### 7.1 Methods

We used a two-level strategy to search for studies evaluating the effect of wildfire smoke exposure on five health outcomes: all-cause mortality, respiratory-related ED visits and hospitalizations (separately), and cardiovascular-related ED visits and hospitalizations (separately). First, we conducted a literature search of the National Library of Medicine’s PubMed database. The full search string is provided in the Supplemental Information. Second, we identified additional articles using search techniques like backward and forward citation chasing, and included references cited in previous systematic and narrative reviews on wildfires and health (namely, refs.^22, 32, 61–65, 71, 104^).

We included studies meeting the following criteria: (a) published in a peer-reviewed journal; (b) human subject studies of the general population; and (c) studies of exposures to wildfire smoke. We excluded articles that did not generate original effect estimates, that evaluated the effects of smoke from other types of fires (e.g., mine fires), and that documented chronic health effects of wildfire smoke pollution.

Titles and abstracts were screened by CFG using the Covidence online platform where duplicate papers were automatically removed. CFG reviewed the full texts of all potentially-eligible studies and determined inclusion. CFG, SHN, and MB reviewed included studies.

#### 7.1.1 Statistical analysis

We extracted risk estimates from all studies and their corresponding confidence intervals for the association between a measure of wildfire smoke exposure and the outcome of interest. When studies presented cumulative and contemporaneous effects of wildfire smoke exposure on health, the contemporaneous (i.e., same-day or lag 0) effect was selected. In the case that lagged effects were presented and/or modeled separately, we extracted the same-day effect estimate. Study-specific estimates were pooled using a random-effects maximum likelihood (REML) estimation approach implemented using the ‘metafor’ package (version 2)^105^ in R (version 4.2.2).^106^ We generated study-specific pooled estimates using REML in the case that studies provided multiple effect estimates (e.g., one estimate per geographic unit).

Between-study heterogeneity was assessed using the *I*^2^ index, which is estimated as the fraction of total heterogeneity explained by between-study heterogeneity. Publication bias was assessed using the Egger’s regression test for funnel plot asymmetry, which tests for the presence of a relationship between observed effect sizes and standard errors.

### 7.2 Results

#### 7.2.1 Description of included studies

The search yielded 1,283 articles (Figures S2–S4). After applying exclusion/inclusion criteria, 153 were eligible for full-text review. Of these, studies were excluded because they did not evaluate the appropriate outcome, were not of the general population (e.g., included older adults or children only), did not generate original effect estimates, had an inadequate study design (e.g., inadequate control for confounders), did not plausibly estimate the effect of wildfire-specific pollution (e.g., estimated the effect of wildfire smoke plumes, event studies), or were not wildfires (e.g., mine fires). For wildfire specific PM_2.5_, we included 8 studies in our meta-analysis of all-cause mortality, 9 for respiratory hospitalizations, 9 for cardiovascular hospitalizations, 5 for respiratory ED visits, and 4 for cardiovascular ED visits. These studies are summarized in Tables S2–6.

#### 7.2.2 Meta-analysis

Figure 3 summarizes included studies and pooled estimates. Same day all-cause mortality increased by 0.15% (95% CI, 0.01%–0.28%) per 1 *μgm*^−3^ increase in wildfire specific PM_2.5_. There were robust positive associations between wildfire PM_2.5_ and same-day respiratory outcomes: respiratory hospitalizations increased by 0.31% (95% CI, 0.08%–0.54%) and respiratory ED visits increased by 0.41% (95% CI, 0.23%–0.60%) per additional *μgm*^−3^ increase in ambient wildfire smoke PM_2.5_. We found a non-statistically-significant 0.04% (95% −0.02%–0.11%) increase in same-day cardiovascular hospitalizations and no meaningful change in same-day cardiovascular ED visits (−0.06%, 95% −0.26%–0.14%) per additional 1 *μgm*^−3^ increase in ambient wildfire smoke PM_2.5_. For all outcomes except respiratory hospitalizations and cardiovascular ED visits, there was evidence of heterogeneity in effects across studies (i.e., Q-statistic P<0.05). Egger’s tests did not indicate evidence of publication bias for any outcome (P>0.05) (see Figure S2 for funnel plots).

**Figure 3:**
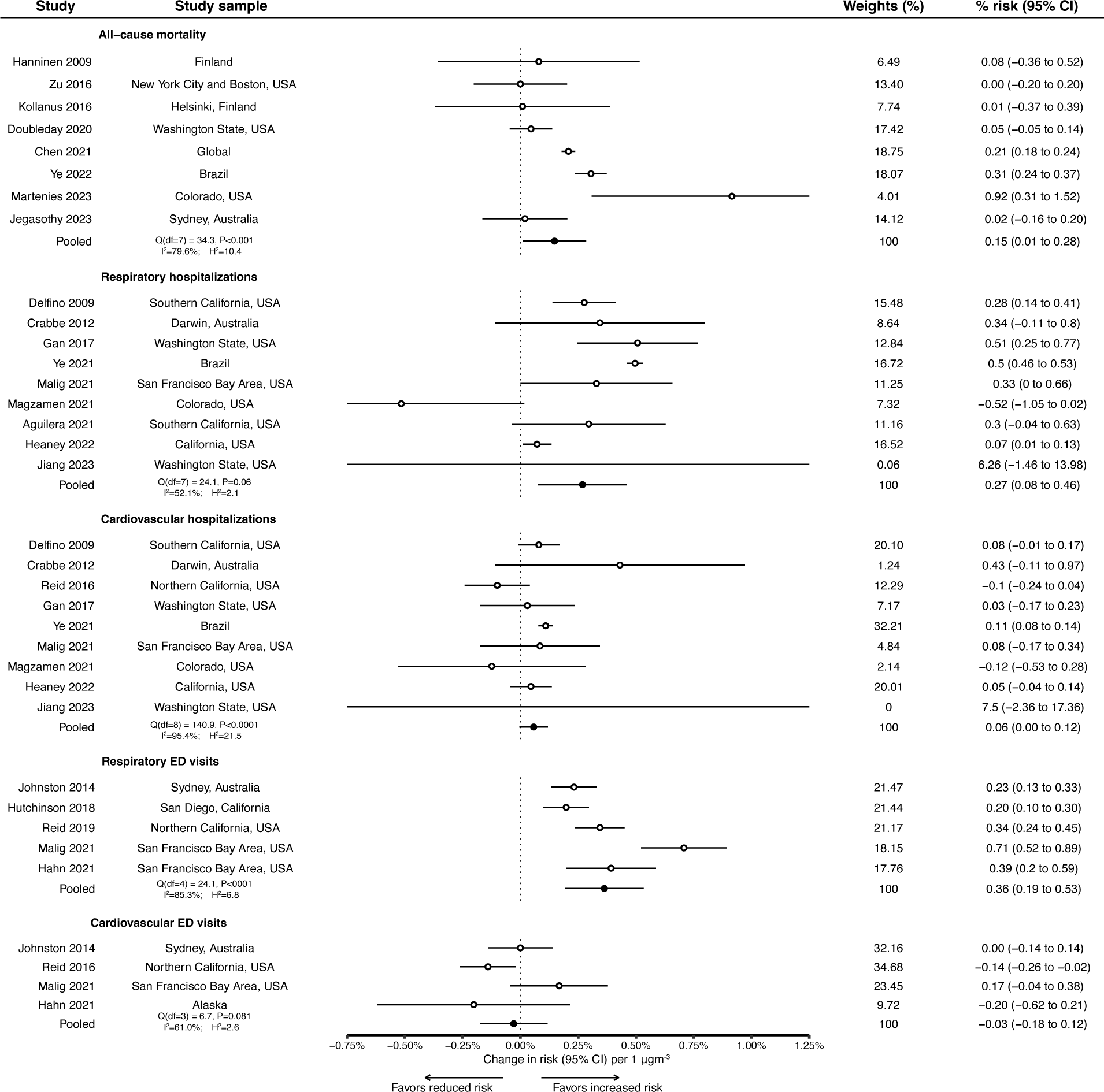
Meta-analysis of the associations between ambient wildfire-specific fine particulate matter and same-day health outcomes per 1 *μgm*^−3^. Pooled responses are derived from random effects meta-analysis estimated via restricted maximum likelihood.

### 7.3 Limitations

There are limitations of our analysis worth discussing. First, as additional studies are published that meet high empirical standards, this meta-analysis should be updated. Second, while we analyze respiratory- and cardiovascular-specific outcomes, these categories are still broad and there could be meaningful cause-specific heterogeneity in responses (i.e., respiratory tract infections vs. chronic respiratory disorders). Third, we only extracted estimates from the general population; further investigation into heterogeneous effects across subpopulations (i.e., sex, age, or socioeconomic and demographic characteristics) is warranted.

Fourth, we extracted same-day effects, which were the most common outcomes reported in the literature, and did not include lagged or cumulative effects as they were not sufficiently consistently reported. However, as more work emerges, including lagged and cumulative effects in meta-analyses will be critical in order to completely quantify the impacts of wildfire smoke on health.

## 8 KNOWLEDGE GAPS

One central knowledge gap in our understanding of the health effects of wildfires is, “How different is pollution from wildfires from other sources in ways that matter for human health?”

The relative composition and toxicity of wildfire smoke to air pollution from other sources remains poorly understood. To investigate the toxicological profile of wildfire pollution compared to other common sources of pollution, future work could aim to answer the question of whether a given unit of PM_2.5_ from wildfire smoke more toxic for human health than the average unit of PM_2.5_ from other sources. However, the answer will likely vary across contexts (e.g., forest type, soil type, whether buildings burned) and exposure pathways. Indeed, evidence suggests that the toxicity of wildfire smoke is related to the distance the smoke traveled since emission. Ability to understand and predict the toxicity of smoke emitted from specific fires could shape long-term and immediate public health response and fire management.

Wildfires are episodic, i.e., do not steadily emit pollution at a fixed level, which distinguishes them from other sources of pollution like transportation and industry. Yet we do not know whether varying patterns of exposure to wildfire smoke have differential impacts on human health. For example, wildfires may lead to ground-level PM_2.5_upwards of 100 *μgm*^−3^ for only a day. But other times, wildfire-specific ambient PM_2.5_might remain 10 *μgm*^−3^ but persist for ten days. The cumulative dose in both scenarios would be 100 *μgm*^−3^, but whether the impacts on human health would differ is not clear.

Another theme for future investigation is the extent to which human behavioral responses to ambient wildfire smoke shape human health outcomes. Given the salience of wildfire smoke and that existing public health strategies rely on individuals undertaking self-protective behaviors when thick smoke is present, understanding the extent to which these behaviors actually protect health and alter health responses is critical for informing future resource allocation and policy. As the above non-linear ED results suggest, the mixed evidence on the health impacts of wildfires may be partially explained by limited changes in behavior on moderate smoke days, but protective behavior on high smoke days. Further, studies focused on cumulative effects of wildfire smoke on healthcare utilization could be averaging the positive and negative effects of wildfire smoke at different exposure levels, leading to null results.

The long-term health impacts of wildfire smoke exposures also remain poorly understood.^107, 108^ These analyses are empirically challenging, as discussed in Section 6, because disentangling variations in wildfire smoke exposure from factors correlated with health outcomes is increasingly difficult over longer time periods. However, as wildfires are a seasonal exposure that can contribute up to half of all ambient air pollution in some regions,^42^ they are worth quantifying to understand optimal investments in control measures.

There are a range of other uncertainties in our understanding of the health impacts of wildfires, including, but certainly not limited to, the imperfect measurement of wildfire smoke pollution discussed in previous sections, the extent to which personal exposures (including indoor exposures) deviate from the ambient exposures used in most health studies, the extent to which prescribed burns are harmful for health and how these negative health impacts are offset by the reduced risk of more harmful fires in the future, the extent to which variations in species present in wildfire smoke are captured in existing environmental epidemiological studies of ambient wildfire-specific pollution by being correlated with total PM_2.5_, the potentially synergistic negative health impacts of hot and smokey days, the extent to which wildfire smoke waves are worse than periodic single day episodes (e.g., four consecutive days vs. four non-consecutive days in the same month) due to biological or behavioral change, and how the salience of wildfires varies across contexts and the extent to which these differences affect health outcomes.

## 9 STRATEGIES FOR ADDRESSING HEALTH EFFECTS OF WILDFIRE SMOKE

We highlight three broad solution areas for addressing the human health effects of wildfires and smoke: those that aim to limit (a) the ignition of health-harming wildfires; (b) the damage from already-ignited wildfires; and (c) the health harm from wildfire smoke.

Addressing the health impacts of wildfires begins with the upstream determinants of wildfire activity. Broadly speaking, the recent increase in wildfire activity in North America has been driven by the combined effect of a century of fire suppression that left an accumulation of fuels, a warming climate that has made these fuels drier and more flammable, and increased human activity in the wildland-urban interface that have made ignitions more likely. Thus reducing the likelihood of future extreme wildfires and the smoke they cause will require addressing these interacting factors. Such efforts are critical but not be easy: global climate change must be slowed or reversed, incentives to build houses in the wildland-urban interface reduced, and a century of accumulated fuels will need to be cleared from fire-prone areas using a variety of fuels management techniques. At scale, such efforts will likely take decades or longer to fully take effect.

When fires are already ignited, difficult decisions must be made as to how to manage them. Historically, efforts have focused on quickly suppressing fires, with suppression activity and costs focused on preventing incursion of fires into human inhabited area. Wildfires near urban areas undoubtedly threaten lives, but fires distant from inhabited areas can generate large downwind smoke exposures that also threaten lives; these more distant fires may receive less suppression effort, even if they are potentially more costly from a public health perspective.^109^ There is also growing recognition that low intensity fire, when left to burn or — in the case of prescribed fire — purposefully ignited, plays a critical ecological function and can reduce the likelihood of future extreme wildfire. Formally quantifying these trade-offs is a critical area for future work.

Given that increasingly extreme wildfire activity and smoke generation is, unfortunately, likely in the near term, efforts to protect public health in the face of growing exposures will be critical. One such effort will be ensuring that the public is informed of when wildfire smoke exposures are expected and how to protect themselves from smoke. Systems can be developed that forecast wildfire activity (e.g.,^110, 111^) and that can be used to warn the public of imminent smoke exposures to encourage health-protective behaviors that limit exposure (e.g., running air purifiers, wearing respiratory protection, leaving the area).^12, 104, 112, 113^ However, evidence suggests that information alone is unlikely to be sufficient for self-protection,^114^ and thus communities will likely need to take direct preventative actions (e.g., subsidy or short-term rentals of air purifiers, access to clean air spaces, shelters in cleaner-air regions) targeted at those most vulnerable, including pregnant individuals, young children with asthma, older adults with chronic lung disease, and outdoor laborers. Physicians can play a role in facilitating preventative action, especially by encouraging at-risk patients to stay at home, run air filters, and take other actions to protect themselves from wildfire smoke, including by prophylactic pre-filling of relevant prescriptions and increasing telemedicine opportunities. Conditional on a given level of smoke exposure, increased health care access can also help mitigate the severity of health impacts.

## 10 CONCLUSIONS

Wildfires are projected to increase in frequency and size in many regions globally because of a combination of climatic and human behavioral factors; the impacts of these fires on air quality and human health are also likely to grow. While accumulating evidence makes it clear that inhaled wildfire smoke negatively impacts human health, it is also increasingly clear that wildfire smoke is different from pollution from other sources in ways that likely matter for human health. For example, wildfire smoke may have a different toxicological profile from pollution from other sources, wildfires produce different patterns of exposure, and the salience of wildfire smoke can induce behavioral changes that alter health impacts, at least in some contexts. Analyses of the health effects of wildfire smoke exposures, including when distant from fires, should take advantage of recently-developed smoke-specific ambient PM_2.5_ datasets and the replicable approaches employed in the production of these data. Future work should aim to leverage and understand the unique dynamics of wildfire smoke: (a) temporal variation in ambient wildfire smoke concentrations is frequently idiosyncratic, enabling causal interpretations when appropriately controlling for area characteristics and time trends, (b) healthcare utilization may respond nonlinearly to increasing pollution levels due to a combination of individual-level pathophysiological impacts and changing behaviors at the individual and population level, and (c) healthcare utilization and health impacts can have varied lagged effects according to the outcome and location of interest. Better understanding these dynamics will be critical for understanding and mitigating the health impacts of wildfires in a changing climate.

## DATA AND CODE AVAILABILITY

Relevant code will be made publicly available upon publication via GitHub. Emergency department data supporting this research are not available for sharing with the public or research community. To request California epidemiological data, researchers may contact the California Department of Healthcare Access and Information at https://hcai.ca.gov/; access fees may apply.

## ACKNOWLEDGMENTS

The authors are grateful to members of the ECHO Lab at Stanford University and Dr. Alexandra K. Heaney for their thoughtful comments.

## SUPPLEMENTAL INFORMATION

Supplemental Information can be downloaded here.

