## Supplemental Information for "Health effects of wildfires"

1 Doerr School of Sustainability, Stanford University, Stanford, USA, 94305;
2 Center on Food Security and the Environment, Stanford University, Stanford, USA, 94305;
3 Department of Medicine, Stanford University, Stanford, USA, 94305;
4 Sean N Parker Center for Allergy and Asthma Research, Stanford University, Stanford, USA, 94305
5 Center for Community Health Impact, The University of Texas Health Science Center at Houston School of Public Health, El Paso, Texas, USA, 79902;
6 National Bureau of Economic Research, Boston, USA, 02138;
7 Environmental Health Sciences, Harvard University, Cambridge, USA, 02115;
^⋄^ These authors contributed equally**.**

^+^ Corresponding author.

**Table S1. Summary of available data sources and approaches for identifying wildfire-specific ambient air pollution concentrations.**

| **Product / Data** | **Approach** | **Inputs** | **Geographic coverage** | **Temporal resolution** | **Spatial resolution** | **Strengths** | **Weaknesses** | **Reference** |
| --- | --- | --- | --- | --- | --- | --- | --- | --- |
| **Smoke PM2.5 datasets** | Statistical modeling | Smoke plumes, station data, other satellite-derived measures (aerosol optical depth), meteorological information, elevation, surface data | Broad | High |  | High spatial and temporal resolution, wide spatial coverage, bounded by observations. Not very computationally costly. Publicly-available, low barrier to entry needed to use. | Reliant on smoke plumes. No perfect measure of smoke PM2.5 on which to train models. Does not directly account for atmospheric chemistry. |  |
| *Examples* |  |  |  |  |  |  |  |  |
| *O'Dell et al. 2019* |  |  | CONUS | Daily, 2006-2016 | 15km x 15 km grid |  |  | (1) |
| *Childs et al. 2022* |  |  | Contiguous US (CONUS) | Daily | 10km x 10km grid |  |  | (2) |
| *Aguilera et al. 2023* |  |  | California | Daily, 2006-2020 | Zip code |  |  | (3) |
| **Total PM2.5 datasets** | Statistical modeling (and hybrid CTMs) | Station data, other satellite-derived measures (aerosol optical depth), meteorological information, elevation, surface data, may also include smoke plumes, chemical transport models | High | High |  | Same as above. | Does not separate wildfire smoke PM from PM from other sources. Same as above. |  |
| *Examples* |  |  |  |  |  |  |  |  |
| *Di et al. 2019* |  |  | CONUS | Daily, 2000-2015 | 1km x 1km grid |  |  | (4) |
| *Reid et al. 2019* |  |  | western US | Daily, 2008-2018 (without CMAQ), 2008-2016 (with CMAQ) | County / census tract / zip code |  |  | (5) |
| *O'Dell et al. 2019* |  |  | CONUS | Daily, 2006-2016 | 15km x 15 km grid |  |  | (1) |
| **Other gridded data sources** |  |  |  |  |  |  |  |  |
| Chemical transport models |  |  |  |  |  | Sophisticated models that can account for complex atmospheric chemistry. | Extremely computationally intensive, leading to difficulties in generating highly-spatially and temporally-resolved estimates. |  |
| *van Donkelaar* | Weighted combination of statistical and chemical transport models | Station data, other satellite-derived measures (aerosol optical depth), meteorological information, elevation, surface data, may also include smoke plumes, chemical transport models | Global | Monthly, 1998-2019 | 2 degree x 2.5 degree grid | Total PM2.5. Highly sophisticated model with global coverage over a long period of time. | Note: Estimates are subject to change as models are updated. Higher spatial resolution datasets are available, but some inputs are only available at higher resolutions and therefore fine resolution gradients may be difficult to interpret. | (6) |
| *van Donkelaar* | Weighted combination of statistical and chemical transport models | Station data, other satellite-derived measures (aerosol optical depth), meteorological information, elevation, surface data, may also include smoke plumes, chemical transport models | North America | Monthly |  | Speciated PM2.5 |  | (6) |
| **Station data** | Monitoring stations | Monitoring stations | Limited | High |  | Best measure of actual air pollution concentrations and can provide total and speciated data. | Poor geographic coverage, only representative of immediate area, subject to data missingness. Still doesn't measure WF smoke PM directly. | (7) |
| **Smoke plume data** | NOAA analyst drawings of where wildfire smoke is in the air | Satellite imagery | North America | Daily |  | Defines the locations of wildfire smoke in easily-accessible shape files on a daily basis over North America. | Limited temporally coverage, are subject to human error, and limited by the ability of people to distinguish where a plume starts and ends, plumes do not identify height of smoke, plumes only as good as satellite imagery so limited by cloud cover. Don’t distinguish between types of smoke plumes (not just wildfires, but also agricultural fires). | (8) |

Links to data sources

1. <https://doi.org/10.1021/acs.est.8b05430>
2. <https://pubs.acs.org/doi/full/10.1021/acs.est.2c02934>
3. <https://www.sciencedirect.com/science/article/pii/S0160412022006468>
4. <https://doi.org/10.1016/j.envint.2019.104909>
5. <https://www.nature.com/articles/s41597-021-00891-1>
6. This group has produced many air pollution estimates, which are well-described and freely-available from repositories accessible on this website: <https://sites.wustl.edu/acag/datasets/surface-pm2-5/>
7. <https://www.epa.gov/outdoor-air-quality-data>
8. <https://www.ospo.noaa.gov/Products/land/hms.html>

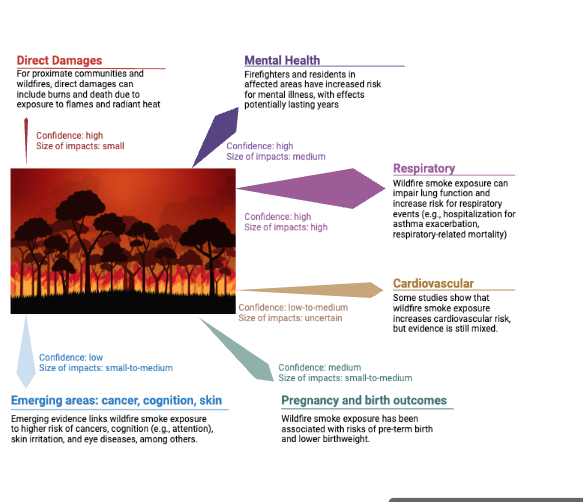

**Figure S1. Summary of the health impacts of wildfires.**

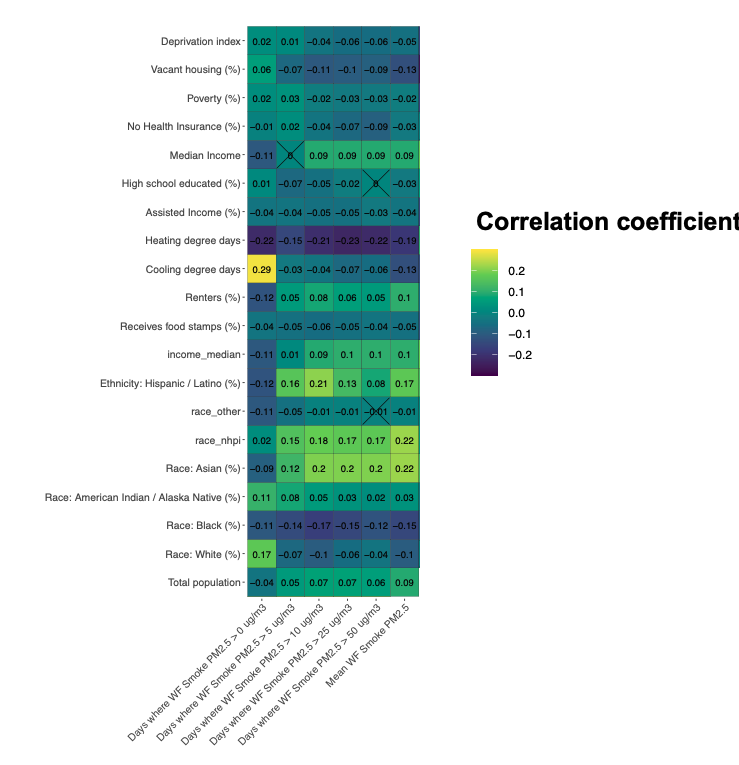

**Figure S2. Cross-sectional correlations between Zip code level ambient wildfire-specific air pollution parameters (from Childs et al. 2022) and socio-economic and demographic characteristics from the American Community Survey averages from 2018-2020.**

**PUBMED Search String**

**("forest fire" OR "bush fire" OR "bushfire" OR "wildfire" OR "wildland fire" OR "landscape fire" OR "prescribed fire" OR "mountain fire" OR "brushfire" OR "brush fire" OR "vegetation fire") AND ("health" OR "public health" OR "human health" OR "population health" OR "community health" OR "mortality" OR "death" OR "morbidity" OR "hospital admission" OR "hospitalisation" OR "hospitalization" OR "emergency department visit" OR "emergency department presentation" OR "ED visit" OR "ambulance" or "respiratory" or "cardiovascular" or "symptom" or "lung function" or "mental health" or "blood pressure")**

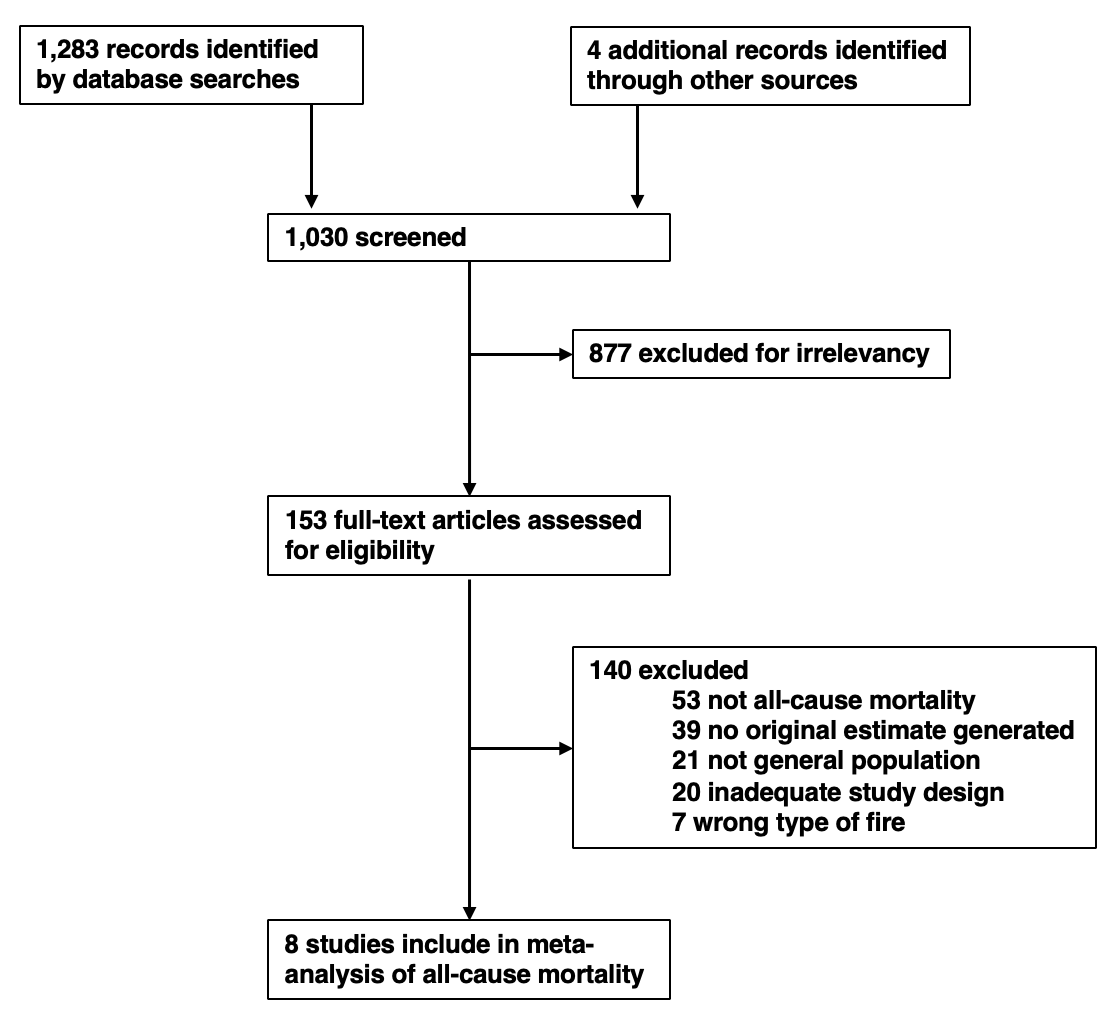

**Figure S3. Flow diagram of the exclusion and inclusion of studies for meta-analysis of all-cause mortality and wildfire-specific air pollution.**

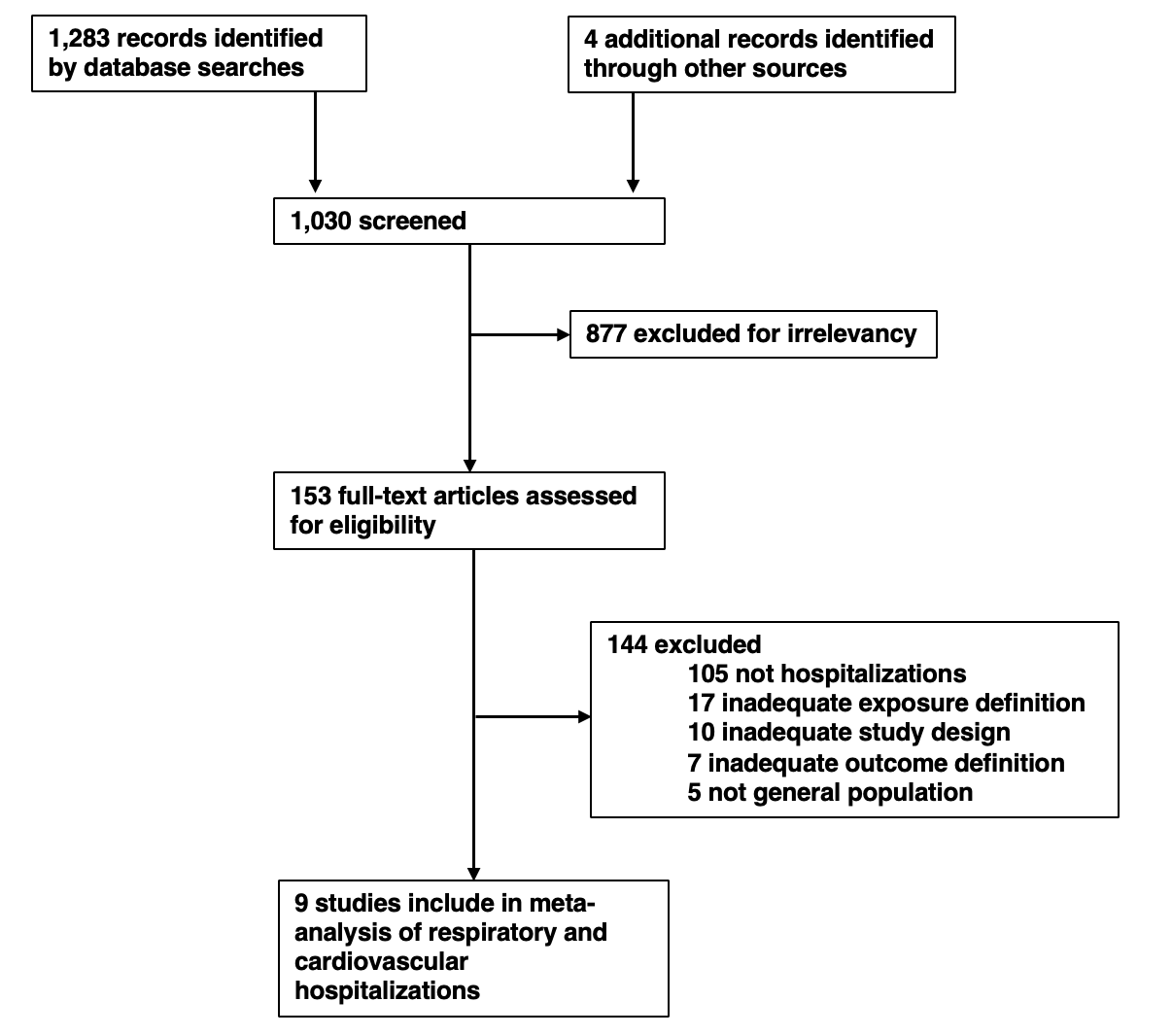

**Figure S4. Flow diagram of the exclusion and inclusion of studies for meta-analysis of hospitalizations and wildfire-specific air pollution.**
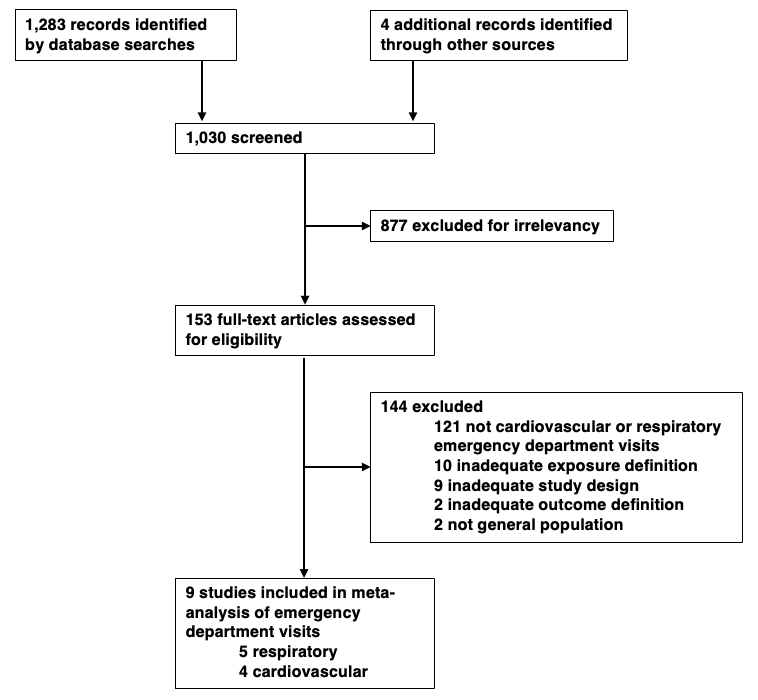

**Figure S5. Flow diagram of the exclusion and inclusion of studies for meta-analysis of emergency department visits and wildfire-specific air pollution.**

**Table S2. Studies included in meta-analysis of ambient wildfire PM2.5 and all-cause mortality**

| **Study** | **Sample period** | **Sample region** | **Geographic unit** | **Exposure definition** | **Outcome Definition** | **Study Design** | **Controls** | **Model formulation** | **Lags** |
| --- | --- | --- | --- | --- | --- | --- | --- | --- | --- |
| Hanninen 2009 | August 26 - September 8, 2002 | Finland | Province (n=11) | Ambient PM2.5; whole period considered wildfire | All deaths | Time series | linear term for time trend; tested dummy for weekday | poisson time series regression | 0-4 |
| Kollanus 2016 | 2001-2010 | Helsinki metro area, Finland | City (n=1) | Ambient PM2.5; wildfire days based on 24-hr >25; 24-hr background >20; NAAPS model >1 (n=72) | Death due to natural causes (ICD-10 A00-R99) | Time series | Mean T, relative humidity, weekly influenza counts (dummied), pollen count, public holiday; triple interaction between year, month, day of week | poisson time series regression (with interaction, similar to time-stratified case-crossover) | 0-4 |
| Zu 2016 | July 7–16, 2002, with reference data from 2001 and 2003 | New York City and Boston, USA | City (n=2) | Ambient PM2.5 | Deaths due to natural causes | Time series | apparent temperature, week of the month, weekend, holiday, year; random effect for city | negative binomial time series regression | 0-5 |
| Doubleday 2020 | June 1 to September 30, yearly 2006-2017 | Washington, USA | Individual level home address coordinates (n=31,719 with exposure variation) | Smoke day; when PM2.5 > 20.4; for 9-20.4, 1) The day must be part of an event in which at least 2 of 3 consecutive days are greater than 9 μg/m3; 2) One of the days in the 3-day event window must be greater than 15 μg/m3; 3) For urban areas (Seattle, Tacoma, Spokane), at least 50% of the air monitors in those areas must be greater than 9 μg/m3 | Non-traumatic cause of death (ICD-10 A01-V99) | Case-crossover | Humidex, time-stratified referent sampling | Individual level conditional logistic regression where strata are the same day of the week, month, and year of death. | 0-4 / 0-1 |
| Chen 2021 | 2000-2016 | Global (n=43 countries) | City (n=749) | Wildfire PM2.5 | Death due to natural causes (ICD-9 0-799; ICD-10 A00-R99) | Time series | Seasonal and long-term trends controlled using natural subic spline of time with 7 degrees of freedom per year; moving average of temperature; relative humidity; day of week. | City-level time series quasi-Poisson regression aggregated via random-effects meta-analysis | 0-7 |
| Ye 2022 | 2000-2016 | Brazil | Regions (n=510) | Wildfire PM2.5 | All deaths | Time series | spline mean T, spline relative humidity, day of week, holidays, seasonality and long-term trends via spline with 7 df per year | district-level time series quasi-Poisson regression distributed-lag non-linear models aggregated via random-effects meta-analysis | 0-14 |
| Jegasothy 2023 | March 1, 2010 - February 29, 2020 | Sydney, Australia | Subregion (n=3) | Wildfire PM2.5 | All deaths | Time series |  | quasi-Poisson regression distributed-lag non-linear models | 0-7 |
| Martenies 2023 | 2010-2020 | Colorado, USA | County (n=11) | WF Smoke Day based on HMS Plumes; standardized using median population-weighted average ambient PM2.5 on smoke vs. non-smoke days | All deaths | Time series | year, day of week, PM2.5, ozone, ambient T, spline day of year | county-level time series negative binomial generalized additive models aggregated via random-effects meta-analysis | 0-2 |

A All studies are at the daily level.

B All studies were included in the meta-analysis.

C Study did not present effect estimate of interest in a table, so the point estimate and confidence intervals were extracted visually using Plot Digitizer.

| Augusto 2020 | October 2017 | Portugal | District (n=14) | Ambient PM10; smoky, dust-free days identified via satellite imagery (n=14) | Death due to natural causes (ICD-10 A00-R99) | Time series | NO2, SO2, O3, max T | District-level time series Poisson regression aggregated via random-effects meta-analysis | None |
| --- | --- | --- | --- | --- | --- | --- | --- | --- | --- |

**Table S3. Studies included in meta-analysis of ambient wildfire PM2.5 and respiratory-related hospitalizations**

| **Study** | **Sample period** | **Sample region** | **Geographic unit** | **Exposure definition** | **Outcome Definition** | **Study Design** | **Controls** | **Model formulation** | **Lags** |
| --- | --- | --- | --- | --- | --- | --- | --- | --- | --- |
| Delfino 2009 | October 21 - October 30, 2003 | Southern California, USA | Zipcode (n~566) | Two-day average ambient PM2.5 during wildfire smoke period (county dependent) | includes all listed specific respiratory ICD-9 plus 7463 additional admissions for the following ICD-9 codes: 277 (cystic fibrosis), 490 (bronchitis NOS), 494 (bronchiectasis), 495 (extrinsic allergic alveolitis), 506 and 508 (other acute/subacute respiratory conditions due to fumes/vapours, or external agents, not separately analysed because n = 44), 786 (symptoms involving the respiratory system/other chest symptoms). | time series | local pressure gradient, fungal spores, weekend, zip-level population, percentage of non-Caucasians, percentage of females, median household income and age distributions | poisson generalized estimating equations | None |
| Crabbe 2012 | 1993-1998 | Darwin, Australia | City (n=1) | Ambient fine particulate matter (PM2?) | ICD-9 460–519 | time series | minimum temperature, relative humidity, a smoothed spline for seasonal effects (16 DF), ‘date’ for a linear effect over time, day of the week and public and school holidays. | poisson | 0-3 (separately) |
| Gan 2017 | July 1 - October 31, 2012 | Washington State, USA | Individual home address, aggregated to zipcode (n~594) | Three measures: GWR chosen | ICD-9 460–519 | case-crossover | temperature, relative hu8midity, wind speed, precipitation | time-stratified case-crossover; conditional logistic regression with patient-specific strata | 0-5 |
| Reid 2019 | June 20–July 31, 2008 | San Francisco Bay Area, USA | Zipcode (n=753) | Ambient PM2.5 (during WF period)  (average of two days prior to event) | Combined asthma (ICD-9 code 493), COPD (496,491-492), pneumonia (480-486), acute bronchitis (466), and acute respiratory infections (460-465) | time series | daily heat index, natural cubic spline for temporal trend with three DF, zip-level smoking %, % of zip >65 yrs, % zip <5 years, %non-white, median income, day of week FE, holiday FE | poisson generalized estimating equations |  |
| Aguilera 2021 | 1999-2012 (excluding June-August months) | Southern California, USA | Zipcode (n=578) | Ambient WF PM2.5 (sesaonal interpolation approach) | ICD-9 460–519 | time series | flu admissions, weather covariates, day-of-week FE, month-of-year FE, zip code FE, linear time trend | panel fixed effects regression | None |
| Malig 2021 | 10/1–10/18, 2015–2017 | San Francisco Bay Area, USA | County (n=9) | Ambient PM2.5 with dummy variable interaction for WF period (10/9-10/17, 2017) | ICD-10 J00-J99 | time series | weekend FE; sensitivity adjustment for day of month (continuous), year FE; additional sensitivity w/ mean temp | quasi-poisson | 0-2 |
| Ye 2021 | 2000-2015 | Brazil | Municipality (n=1814) | Ambient WF PM2.5 | ICD-10 J00-J99 | time series | natural spline for temperature with three DF, natural spline for relative humidity with three DF, natural spline for time with 7 DF per year, day of week FE, holiday FE. | two-stage regression where municipality-specific quasi-poisson regressions are pooled via random-effects meta-analysis | 0-7 |
| Magzamen 2021 | 2010-2015 | Colorado, USA | Individual home address (n=46,585 cases), aggregated to 15x15km grid | Ambient WF PM2.5 | ICD-9 460–519 | case-crossover | lagged temperature, same-day ozone | time-stratified case-crossover; conditional logistic regression with patient-specific strata | 0-5 |
| Heaney 2022 | 2004-2009 | California, USA | County (n=58) | Smoke event day (cumulative 0-1 day lag WF-specific PM2.5 >=98th percentile) | ICD-9 460–519 | time series | county FE, day of week FE, mean temperature, non-WF PM2.5, month of year FE, year FE |  | 0-14 (separately) |
| Jiang 2023 | August, October, and November 2016 | August, October, and November 2016 | Zipcode (n=160, 351, and 352 per month) | Ambient WF PM2.5 | ICD-10 J00-J99 | time series | 8-hr ozone, temperature, precipitation, wind speed, non-WF PM2.5 | zero-inflated negative binomial regression | None |

**Table S4. Studies included in meta-analysis of ambient wildfire PM2.5 and cardiovascular related hospitalizations**

| **Study** | **Sample period** | **Sample region** | **Geographic unit** | **Exposure definition** | **Outcome Definition** | **Study Design** | **Controls** | **Model formulation** | **Lags** |
| --- | --- | --- | --- | --- | --- | --- | --- | --- | --- |
| Delfino 2009 | October 21 - October 30, 2003 | Southern California, USA | Zip code  (n = ~566) | Two-day average ambient PM2.5 during wildfire smoke period (county dependent) | includes all listed specific cardiovascular ICD-9 codes plus 812 additional admissions for ICD-9 codes 440–459 (diseases of the peripheral circulation) | time series | local pressure gradient, fungal spores, weekend, zip-level population, percentage of non-Caucasians, percentage of females, median household income and age distributions | poisson generalized estimating equations | None |
| Crabbe 2012 | 1993-1998 | Darwin, Australia | City (n=1) | Ambient fine particulate matter (PM2?) | ICD-9 390-459 | time series | minimum temperature, relative humidity, a smoothed spline for seasonal effects (16 DF), ‘date’ for a linear effect over time, day of the week and public and school holidays. | poisson | 0-3 (separately) |
| Reid 2016 | May 6 - September 15, 2008 | Northern California, USA | Zip code (n=781) | two-day moving average Ambient PM2.5 with dummy variable interaction for WF period (June 20-July 31, 2008)  (average of two days prior to event) | Combined ICD-9 ischemic heart disease (IHD) (410–414), cardiac dysrhythmias and conduction disorders (426–427), heart failure (428), cerebrovascular disease (430–435, 437), and hypertension (401–405) | time series | time trend, day of week FE, heat index, median income, % population >65 years, smoking prevalence, ozone |  | None |
| Gan 2017 | July 1 - October 31, 2012 | Washington State, USA | Individual home address, aggregated to zip code  (n = ~594) | Three measures: GWR chosen | ICD-9 390–459 | case-crossover | temperature, relative humidity, wind speed, precipitation | time-stratified case-crossover; conditional logistic regression with patient-specific strata | 0-5 |
| Malig 2021 | 10/1–10/18, 2015–2017 | San Francisco Bay Area, USA | County (n=9) | Ambient PM2.5 with dummy variable interaction for WF period (10/9-10/17, 2017) | ICD-10 I00-I99 | time series | weekend FE; sensitivity adjustment for day of month (continuous), year FE; additional sensitivity w/ mean temp | quasi-poisson | 0-2 |
| Ye 2021 | 2000-2015 | Brazil | Municipality (n=1814) | Ambient WF PM2.5 | ICD-10 J00-J99 | time series | natural spline for temperature with three DF, natural spline for relative humidity with three DF, natural spline for time with 7 DF per year, day of week FE, holiday FE. | two-stage regression where municipality-specific quasi-poisson regressions are pooled via random-effects meta-analysis | 0-7 |
| Magzamen 2021 | 2010-2015 | Colorado, USA | Individual home address (n=46,585 cases), aggregated to 15x15km grid | Ambient WF PM2.5 | ICD-9 390–459 | case-crossover | lagged temperature, same-day ozone | time-stratified case-crossover; conditional logistic regression with patient-specific strata | 0-5 |
| Heaney 2022 ^a^ | 2004-2009 | California, USA | County (n=58) | Smoke event day (WF-specific PM2.5 >=98th percentile) | ICD-9 390–459 | time series | county FE, day of week FE, mean temperature, non-WF PM2.5, month of year FE, year FE | negative binomial panel fixed effects regression | 0-14 (separately) |
| Jiang 2023 | August, October, and November 2016 | Washington State, USA | Zipcode (n=160, 351, and 352 per month) | Ambient WF PM2.5 | ICD-10 I00–I99 | time series | 8-hr ozone, temperature, precipitation, wind speed, non-WF PM2.5 | zero-inflated negative binomial regression | None |

^a^ Effect estimate included in meta-analysis obtained via direct communication with the lead author of this study because the original study did not report standard errors in the supplemental table for lag 0 estimates.

**Table S5. Studies included in meta-analysis of ambient wildfire PM2.5 and respiratory related emergency department visits**

| **Study** | **Sample period** | **Sample region** | **Geographic unit** | **Exposure definition** | **Outcome Definition** | **Study Design** | **Controls** | **Model formulation** | **Lags** |
| --- | --- | --- | --- | --- | --- | --- | --- | --- | --- |
| Johnston 2014 | 1996-2007 | Sydney, Australia | Individual (n=663,333) | Smoke event (n=46) vs. non smoke event ^a^ | ICD-9 460-519  ICD-10 J00-J99 (excluding J95.4 to J95.9), R09.1, R09.8 | Case-crossover | natural cubic splines were fitted to temperature and dew point using four degrees of freedom for temperature splines (same day and lagged) and three degrees of freedom for the dew point splines (same day and lagged), influenza epidemics FE, public holidays FE, and school holidays FE. | Time-stratified case-crossover matched on year, month, and day-of-week estimated using conditional logistic regression | 0-3 |
| Hutchinson 2018 | October 22 - 26, 2007 | San Diego, California | Individual  (n=?) | Ambient PM2.5 during wildfire period | 493, 466, 491, 492, 496, 490, 480-487, 460-464, 277, 494, 495, 786, 506, 508 | Case-crossover | none | Bidirectional symmetric case-crossover matched to two control days equidistant from the event (within two weeks), matched on day of week | none |
| Reid 2019 | June 20–July 31, 2008 | San Francisco Bay Area, USA | Zipcode (n=753) | Predicted ambient PM2.5 (during WF period) (average of two days prior to event) | Combined asthma (ICD-9 code 493), COPD (496,491-492), pneumonia (480-486), acute bronchitis (466), and acute respiratory infections (460-465) | time series | daily heat index, natural cubic spline for temporal trend with three DF, zip-level smoking %, % of zip >65 yrs, % zip <5 years, %non-white, median income, day of week FE, holiday FE | poisson generalized estimating equations | none |
| Hahn 2021 | 2015-2019 wildfire seasons | Alaska | Individual (n=21,263) | Ambient WF PM2.5 | ICD-9: 466, 480-486, 490-494, 496; ICD-10: J12-J18, J20-J22, J40-J45, J47 | Case-crossover | Temperature, relative humidity, | Time-stratified case-crossover; reference = same day of week for duration of wildfire season of the same year, estimated via conditional logistic regression | 0-5 |
| Malig 2021 | 10/1–10/18, 2015–2017 | San Francisco Bay Area, USA | County (n=9) | Ambient PM2.5 with dummy variable interaction for WF period (10/9-10/17, 2017) | ICD-10 I00-I99 | time series | weekend FE; sensitivity adjustment for day of month (continuous), year FE; additional sensitivity w/ mean temp | quasi-poisson | 0-2 |

^a^ Smoke event day mean ambient PM2.5 is 39.1 vs. non-smoke day ambient is 9.9. Observed OR from smoke event vs. non-smoke event was standardized using the difference 39.1 minus 9.9.

**Table S6. Studies included in meta-analysis of ambient wildfire PM2.5 and cardiovascular related emergency department visits**

| **Study** | **Sample period** | **Sample region** | **Geographic unit** | **Exposure definition** | **Outcome Definition** | **Study Design** | **Controls** | **Model formulation** | **Lags** |
| --- | --- | --- | --- | --- | --- | --- | --- | --- | --- |
| Johnston 2014 | 1996-2007 | Sydney, Australia | Individual (n= 368,423) | Smoke event vs. non smoke event | ICD-9 390-459;  ICD-10 I00-I99 (excluding I67.3, I68.0, I88, I97.8, I97.9, I98.0), G45 (excluding G45.3), G46, M30, M31, R58 | Case-crossover | natural cubic splines were fitted to temperature and dew point using four degrees of freedom for temperature splines (same day and lagged) and three degrees of freedom for the dew point splines (same day and lagged), influenza epidemics FE, public holidays FE, and school holidays FE. | Time-stratified case-crossover matched on year, month, and day-of-week estimated using conditional logistic regression | 0-3 |
| Reid 2016 | June 20 - July 31, 2008 | Northern California | Zipcode (n=781) | Predicted ambient PM2.5 during wildfire period (average of two days prior to event) | ischemic heart disease (IHD) (410–414), cardiac dysrhythmias and conduction disorders (426–427), heart failure (428), cerebrovascular disease (430–435, 437), and hypertension (401–405) | Time series | daily heat index, temporal trend modeled with a natural cubic spline with 3 degrees of freedom, estimated ZIP code-level smoking prevalence, % of the ZIP code aged 65 or older, % of the ZIP code aged 5 or younger, % non-White, ZIP code level median income, and day of week and holidays modeled as dummy variables. | poisson generalized estimating equations | none |
| Hahn 2021 | 2015-2019 wildfire seasons | Alaska | Individual (n=5,356) | Ambient WF PM2.5, where PM2.5 >1 standard deviation of long-term monthly mean 2008-2019 and monitor located within 50km of a smuke plume. WF-specific PM2.5 is the difference between long-term monthly mean and daily PM2.5 measured. | 427; I46-I49; 430-438; I60-I63, I65-I69, G45, I23; 410-414; I20-I22, I24-I25; 428; I50 | Case-crossover | Temperature, relative humidity, | Time-stratified case-crossover; reference = same day of week for duration of wildfire season of the same year, estimated via conditional logistic regression | 0-5 |
| Malig 2021 | 10/1–10/18, 2015–2017 | San Francisco Bay Area, USA | County (n=9) | Ambient PM2.5 with dummy variable interaction for WF period (10/9-10/17, 2017) | ICD-10 I00-I99 | time series | weekend FE; sensitivity adjustment for day of month (continuous), year FE; additional sensitivity w/ mean temp | quasi-poisson | 0-2 |

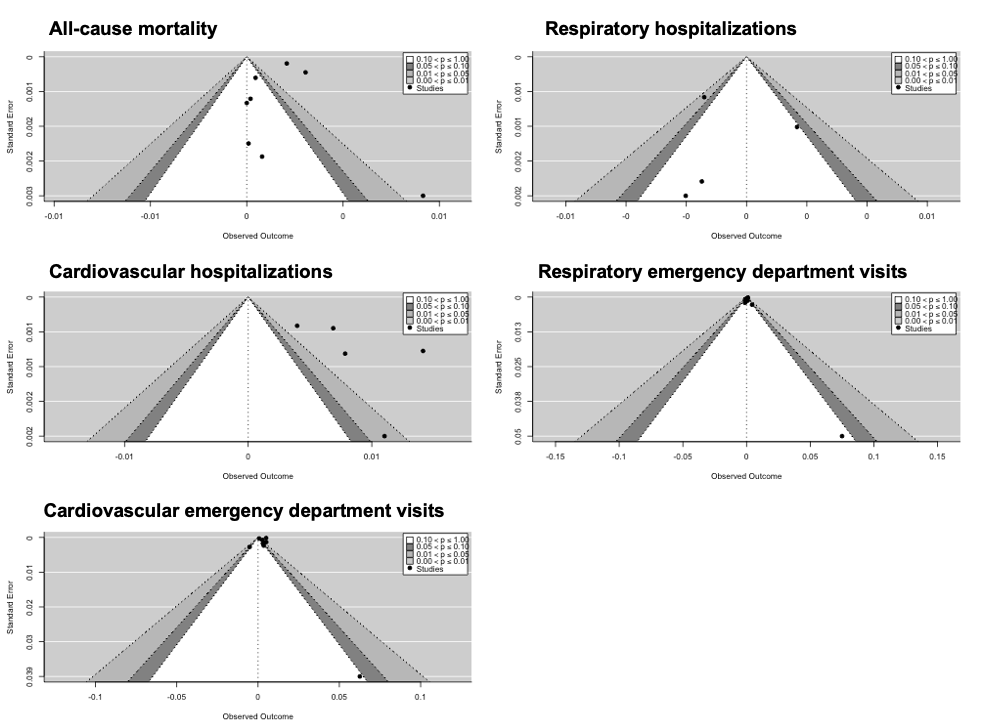

**Figure S6. Funnel plots from meta-analytic summary of the evidence.**
